# The antibody response to SARS-CoV-2 infection persists over at least 8 months in symptomatic patients

**DOI:** 10.1101/2021.02.05.21251219

**Authors:** Riccardo Levi, Leonardo Ubaldi, Chiara Pozzi, Giovanni Angelotti, Maria Teresa Sandri, Elena Azzolini, Michela Salvatici, Victor Savevski, Alberto Mantovani, Maria Rescigno

**Affiliations:** Humanitas University, Department of Biomedical Sciences, Via Rita Levi Montalcini, 20072 Pieve Emanuele, MI, Italy; IRCCS Humanitas Clinical and Research Center, Via Manzoni 56, 20089 Rozzano, MI, Italy; The William Harvey Research Institute, Queen Mary University of London, London, UK

## Abstract

The factors involved in the persistence of antibodies to SARS-CoV-2 are unknown. We evaluated the antibody response to SARS-CoV-2 in personnel from 10 healthcare facilities and its association with individuals’ characteristics and COVID-19 symptoms in an observational study. We enrolled 4735 subjects (corresponding to 80% of all personnel) for three time points over a period of 8-10 months. For each participant, we determined the rate of antibody increase or decrease over time in relation to 93 features analyzed in univariate and multivariate analyses through a machine learning approach. In individuals positive for IgG (≥ 12 AU/mL) at the beginning of the study, we found an increase [*p= 0*.*0002*] in antibody response in symptomatic subjects, particularly with anosmia/dysgeusia (OR 2.75, 95% CI 1.753 – 4.301), in a multivariate logistic regression analysis. This may be linked to the lingering of SARS-CoV-2 in the olfactory bulb.

## Introduction

It is becoming clear that the antibody response to SARS-CoV-2 can last at least 6 months in symptomatic patients ^1^, but it seems to decline in asymptomatics ^2^. Similarly, a reduction of antibody response in asymptomatic individuals was shown in a study with a fewer number of individuals (n = 37) ^3^. The antibody response in COVID-19 patients is associated with the establishment of a memory B cell response which is higher at 6 months ^1^, however, it is not clear whether there are features that correlate with this sustained B cell response. We previously showed that an anti-SARS-CoV-2 serological analysis allowed us to follow the diffusion of the virus within healthcare facilities in areas differently hit by the virus ^4^. At 8-10 months of distance, we analyzed the duration of this antibody response and evaluated whether there were features correlating with maintenance, reduction or increase of the antibody response.

## Results

We analyzed the persistence of the antibody response in healthcare workers that underwent immunological surveillance for SARS-CoV-2 exposure and resulted positive for anti-Spike 1/2 IgG (IgG ≥ 12 AU/mL). Although the test manufacturer considers positive subjects above 15 AU/mL and equivocal those between 12 and 15 (AU/mL), based on our previous publication showing that these two groups behaved very similarly we considered positive everybody ≥ 12 AU/mL^4^. The accrual was on a voluntary basis and did not occur in the symptomatic phase (at around 43 +/- 17 days from COVID-19 assessment when symptomatic). We excluded all of the individuals that became positive over the course of the analysis so to focus only on those individuals that were exposed during the first wave of infection to evaluate the duration of the antibody response. We assessed the correlation of the rate of antibody increase or decrease with the different analyzed features for the first two time points of observation in subjects with IgG ≥ 12 AU/mL. In Tables 1 and 2 are reported the rates for individual classes of features with relative statistical analysis. As shown, females sustained the antibody response better than males (*p = 0*.*01*); similarly non-medical healthcare professionals (specifically, healthcare partner operators) had higher antibody rates (*p = 0*.*0009*). The levels of antibodies increased in hospitals located in the Bergamo area (Castelli and Gavazzeni *p < 0*.*0001*) (Table 1) which was more hit by COVID-19 (37 – 43% of individuals with IgG ≥ 12) ^4^. More important, the IgG rate in individuals which were positive for IgG (IgG ≥ 12 AU/mL; n = 613) at the beginning of the study was increased (*p<0*.*000001*) over time, and this increase was either minor in asymptomatics (n = 91, *p* = 0.00003) and paucisymptomatics (n = 203) or strong in symptomatics (n = 319, *p = 0*.*0006*) (Table 2). This may explain why individuals from hospitals in the Bergamo area or non-medical healthcare professionals had higher levels of antibodies as most of them suffered from symptomatic COVID-19 (59% and 73%, respectively). On the contrary, those that had an intermediate IgG titer (3.8 < IgG < 12 AU/mL considered as negative) displayed all a significant reduction in IgG rate (*p < 0*.*000001*) (Table 1). However, this population is considered as negative for SARS-CoV-2 IgG according to manufacturer. Many symptoms, including fever, cough, muscle pain, asthenia, tachycardia and anosmia/dysgeusia, correlated with an increase of antibodies in the first two time points of observation (Table 2).

**Table 1.**
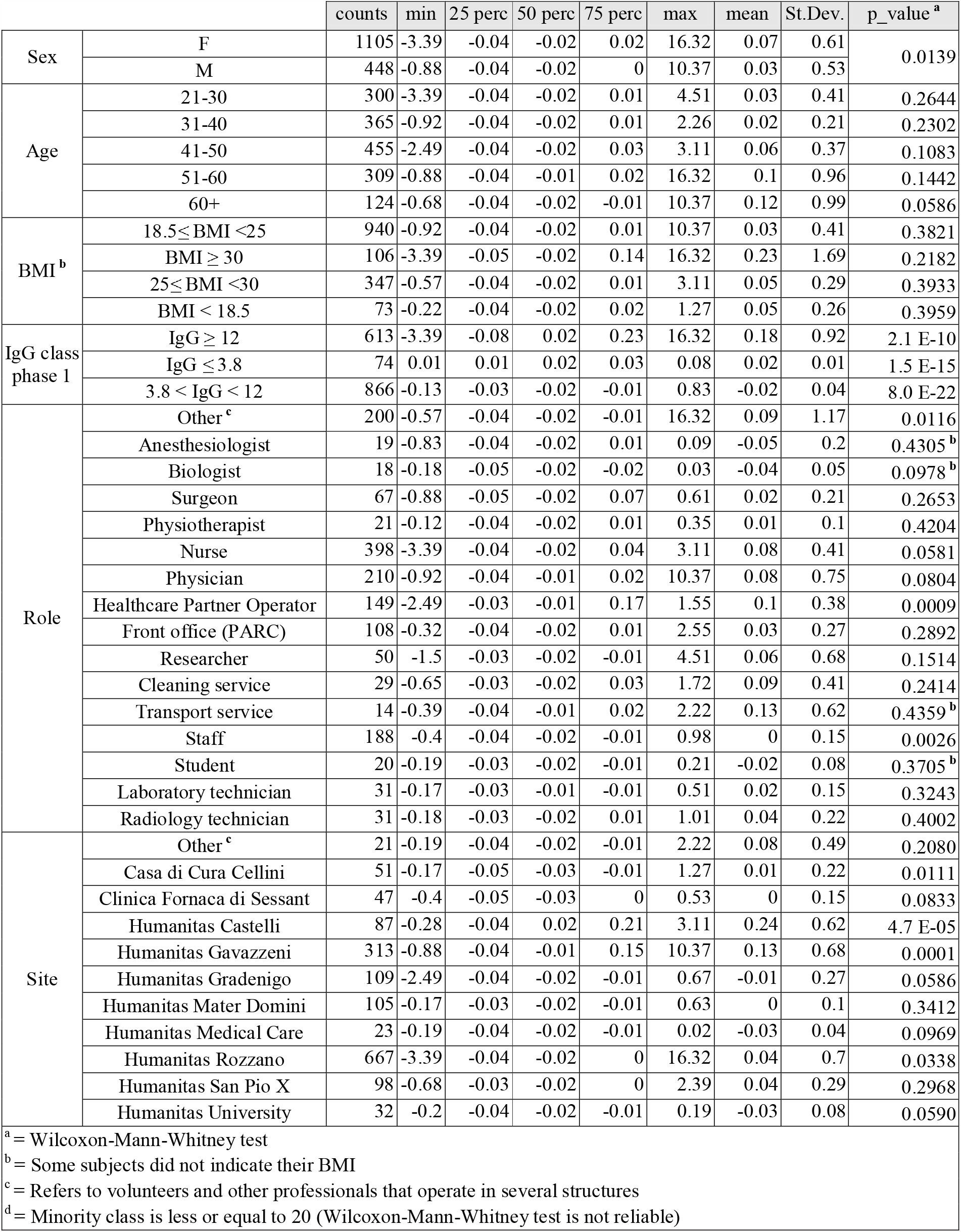
Demographic distribution of antibody rates.

**Table 2.**
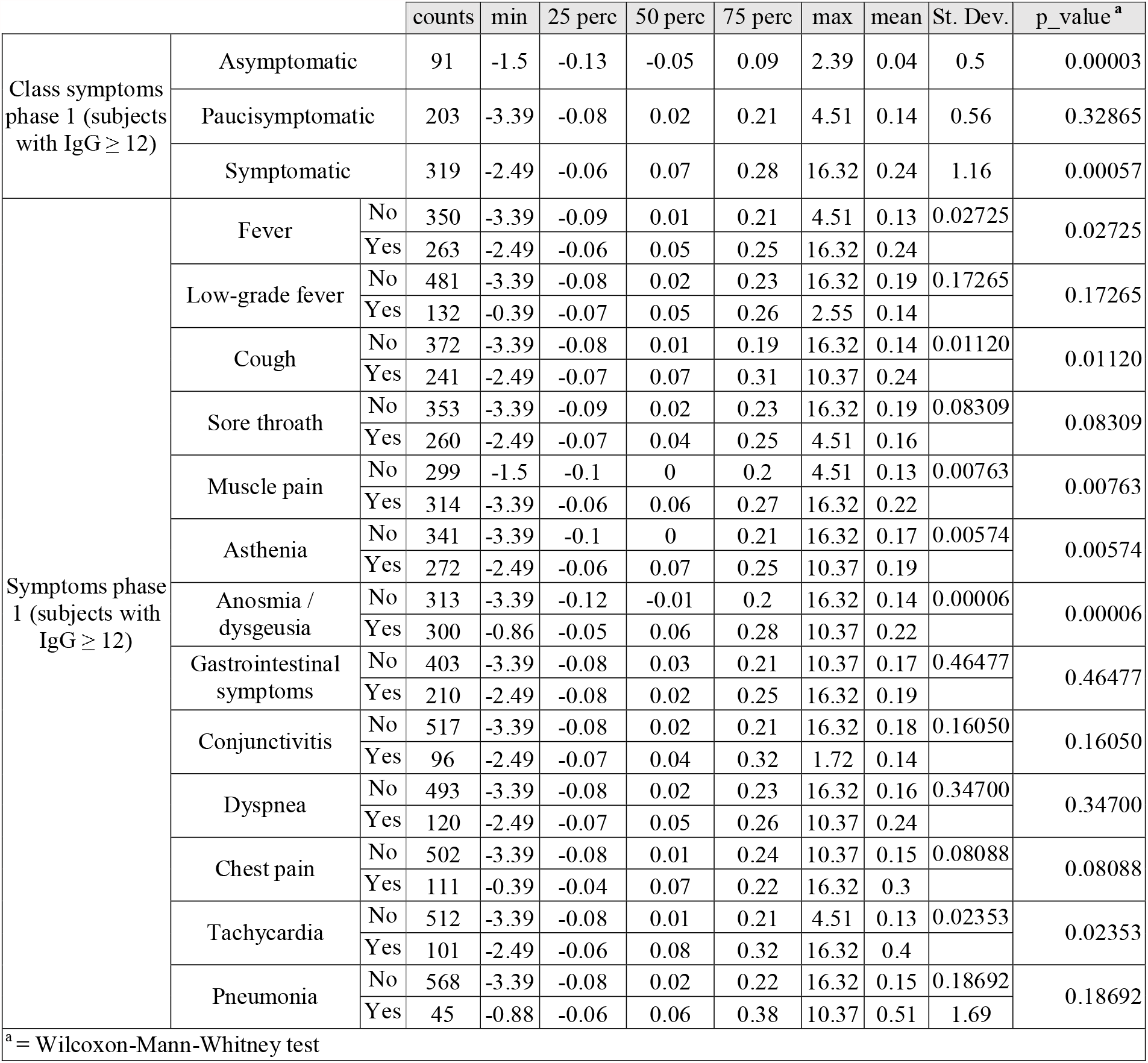
Antibody rates according to symptoms.

As we noticed that the distribution of the rate feature presented a high value of kurtosis (see methods) we restricted the data set to subjects with IgG rates either below the 10^th^ percentile [< - 0.033 (n = 454)] or above the 90^th^ percentile [> 0.005 (n = 445)] to prevent a bias-variance problem in machine learning models. The accuracy of these rates was confirmed by a linear regression analysis. In Figure 1a and 1b are shown the regression diagnostic plots of predicted values against residuals of training and test data according to the threshold (< -0.033 AU/ml*day and > 0.005 AU/ml*day). In Table 3 is shown the Chi-squared analysis for the populations below or above the set threshold rates. We found that as for the previous analysis, males reduced the level of antibodies more than females, even though this difference was no longer statistically significant (*p = 0*.*06*). The levels of antibodies increased in hospitals located in the Bergamo area (Castelli and Gavazzeni: *p = 0*.*0032* and *p = 0*.*0005*, respectively) which was more hit by COVID-19 (37 – 43% of individuals with IgG ≥ 12) and most of the individuals were symptomatic ^4^ while it decreased in Humanitas Rozzano (*p=0*.*0806*) which was less heavily hit (10% of individuals with IgG ≥ 12) and had less symptomatic individuals ^4^ (Table 3). The rate decreased in asymptomatic (65% of subjects fell in the group < -0.033; *p < 0*.*000001*), remained constant in paucisymptomatic and increased in symptomatic individuals (62% of subjects were in the group > 0.005; *p <0*.*000001*) (Table 4). Interestingly, among the different symptoms, fever, cough, muscle pain, asthenia, dyspnea, tachycardia, chest pain and anosmia/dysgeusia all correlated with a higher number of individuals falling into the group with rate > 0.005, indicating that these symptoms were strongly associated with sustained/increased antibody response (*0*.*000001* < *p* < *0*.*05*, Table 4 and Suppl. Fig. 1). Among these, anosmia/dysgeusia was associated with the highest percentage of subjects presenting with increased IgG rate (69%; *p < 0*.*000001*, Table 4 and Suppl. Fig. 1). Having observed differences according to sex, role and site, and since many symptoms are linked, we performed a multivariate statistical analysis based on a supervised machine learning classification approach (see methods). Through a Bayesian hyperparameter optimization algorithm, we assessed that the best machine learning model is a Bagging classifier of 7 logistic regression, which was evaluated both on a training (Accuracy = 76.26; ROC AUC = 76.30; Recall = 81.14) and a test dataset (Accuracy = 72.00; ROC AUC = 72.12; Recall = 81.08), where the training/test split is 80%-20%, stratified by outcome. Classification metrics on training set and test set are comparable, which shows that the model does not present overfitting on training data. In Figure 1c is shown the multivariate logistic regression analysis. We found that the increased rate was associated primarily with anosmia/dysgeusia (regression coefficient=1.0, 95% CI 0.56 – 1.46) and with chest pain (regression coefficient=0.84, 95% CI 0.24 – 1.44), while the decreased rate was associated to subjects with intermediate IgG (3.8 < IgG < 12) (regression coefficient = -1.61, 95% CI -2.03 – -1.0), which may be related to a noise in the instrument testing, and with past neoplasia (regression coefficient = - 1.38, 95% CI -2.4 – -0.37). Interestingly, 54% of subjects with chest pain also presented loss of smell/taste while only 22% of subjects with smell/taste dysfunction also had chest pain, suggesting that IgG increase in the symptomatic population is primarily linked to anosmia and dysgeusia (not shown). In figure 1d are shown the odds ratio relative to figure 1c, which for chest pain is 2.32 (95% CI 1.27 – 4.24), for anosmia/dysgeusia is 2.75 (95% CI 1.75 – 4.30), for subjects with intermediate IgG (3.8 < IgG < 12) is 0.2 (95% CI 0.13 – 0.30) and for subjects with past neoplasia is 0.25 (95% CI 0.09 – 0.69). Overall, these results indicate that although many symptoms are associated with an increase of IgG abundance in the observation time, only anosmia/dysgeusia and chest pain are associated to a higher IgG rate in the multivariate logistic regression analysis. By contrast the population with past neoplasia or intermediate levels of IgG (3.8 < IgG < 12 AU/mL) are the ones that display a reduction in IgG. However, the significance of the reduction in the detection of antibodies in subjects with intermediate levels of IgG remains to be investigated, as this population did not represent early infected individuals because they were all nasopharyngeal swab negative ^4^.

**Figure 1:**
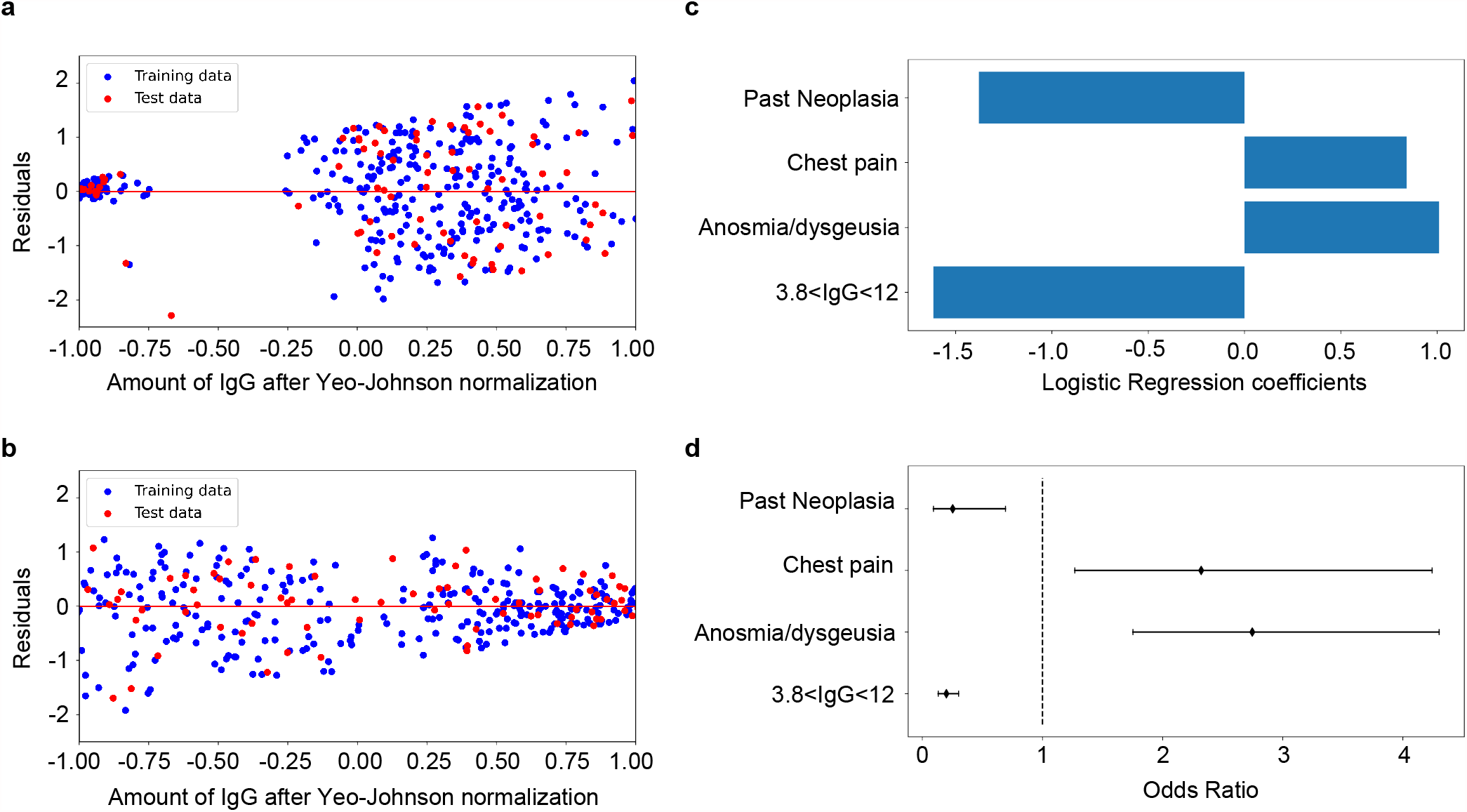
**a**, Dataset < 10^th^ percentile Regression Diagnostic plot of amount of IgG after Yeo-Johnson normalization against residuals of training and test data; **b**, Dataset >90^th^ percentile Regression Diagnostic plot of amount of IgG after Yeo-Johnson normalization against residuals of training and test data; **c**, Barplot with Logistic Regression coefficients for most important features; **d**, Odds ratio of Logistic Regression with confidence intervals (95%) for the most important features.

**Table 3.** Chi-squared analysis of groups < 10^th^ percentile and > 90^th^ percentile.

|  |  | < 10 perc | > 90 perc | p_value |
| --- | --- | --- | --- | --- |
| Sex | F | 321 | 340 | 0.0627 |
|  | M | 133 | 105 |  |
| Age | 21-30 | 93 | 83 | 0.5429 |
|  | 31-40 | 108 | 94 | 0.3804 |
|  | 41-50 | 121 | 142 | 0.0970 |
|  | 51-60 | 88 | 98 | 0.3711 |
|  | 60+ | 44 | 28 | 0.0793 |
| BMI <sup>a</sup> | 18.5≤ BMI <25 | 267 | 255 | 0.6963 |
|  | BMI ≥ 30 | 58 | 72 | 0.1750 |
|  | 25≤ BMI <30 | 106 | 93 | 0.4214 |
|  | BMI < 18.5 | 23 | 25 | 0.8261 |
| Role | Other <sup>b</sup> | 63 | 37 | 0.0109 |
|  | Anesthesiologist | 6 | 7 | 0.9701 |
|  | Biologist | 5 | 2 | 0.4640 |
|  | Surgeon | 25 | 21 | 0.7007 |
|  | Physiotherapist | 6 | 7 | 0.9710 |
|  | Nurse | 113 | 132 | 0.1255 |
|  | Physician | 56 | 61 | 0.6082 |
|  | Healthcare Partner Operator | 38 | 64 | 0.0062 |
|  | Front office (PARC) | 31 | 28 | 0.8494 |
|  | Researcher | 13 | 9 | 0.5485 |
|  | Cleaning service | 7 | 9 | 0.7698 |
|  | Transport Service | 5 | 5 | 0.7747 <sup>c</sup> |
|  | Staff | 69 | 44 | 0.0214 |
|  | Student | 5 | 4 | 0.9759 <sup>c</sup> |
|  | Laboratory Technician | 6 | 7 | 0.9701 |
|  | Radiology Technician | 6 | 8 | 0.7587 |
| Site | Other <sup>b</sup> | 6 | 3 | 0.5223 <sup>c</sup> |
|  | Casa di Cura Cellini | 19 | 8 | 0.0573 |
|  | Clinica Fornaca di Sessant | 18 | 12 | 0.3828 |
|  | Humanitas Castelli | 23 | 47 | 0.0032 |
|  | Humanitas Gavazzeni | 97 | 142 | 0.0005 |
|  | Humanitas Gradenigo | 36 | 22 | 0.0918 |
|  | Humanitas Mater Domini | 20 | 23 | 0.7041 |
|  | Humanitas Medical Care | 8 | 3 | 0.2379 <sup>c</sup> |
|  | Humanitas Rozzano | 190 | 160 | 0.0806 |
|  | Humanitas San Pio X | 27 | 21 | 0.5025 |
|  | Humanitas University | 10 | 4 | 0.1905 <sup>c</sup> |
| <sup>a</sup> = Some subjects did not indicate their BMI |  |  |  |  |
| <sup>b</sup> = Refers to volunteers and other professionals that operate in several structures |  |  |  |  |
| <sup>c</sup> = Minority class is less or equal to 5 (chi-square test is not reliable) |  |  |  |  |

**Table 4.**
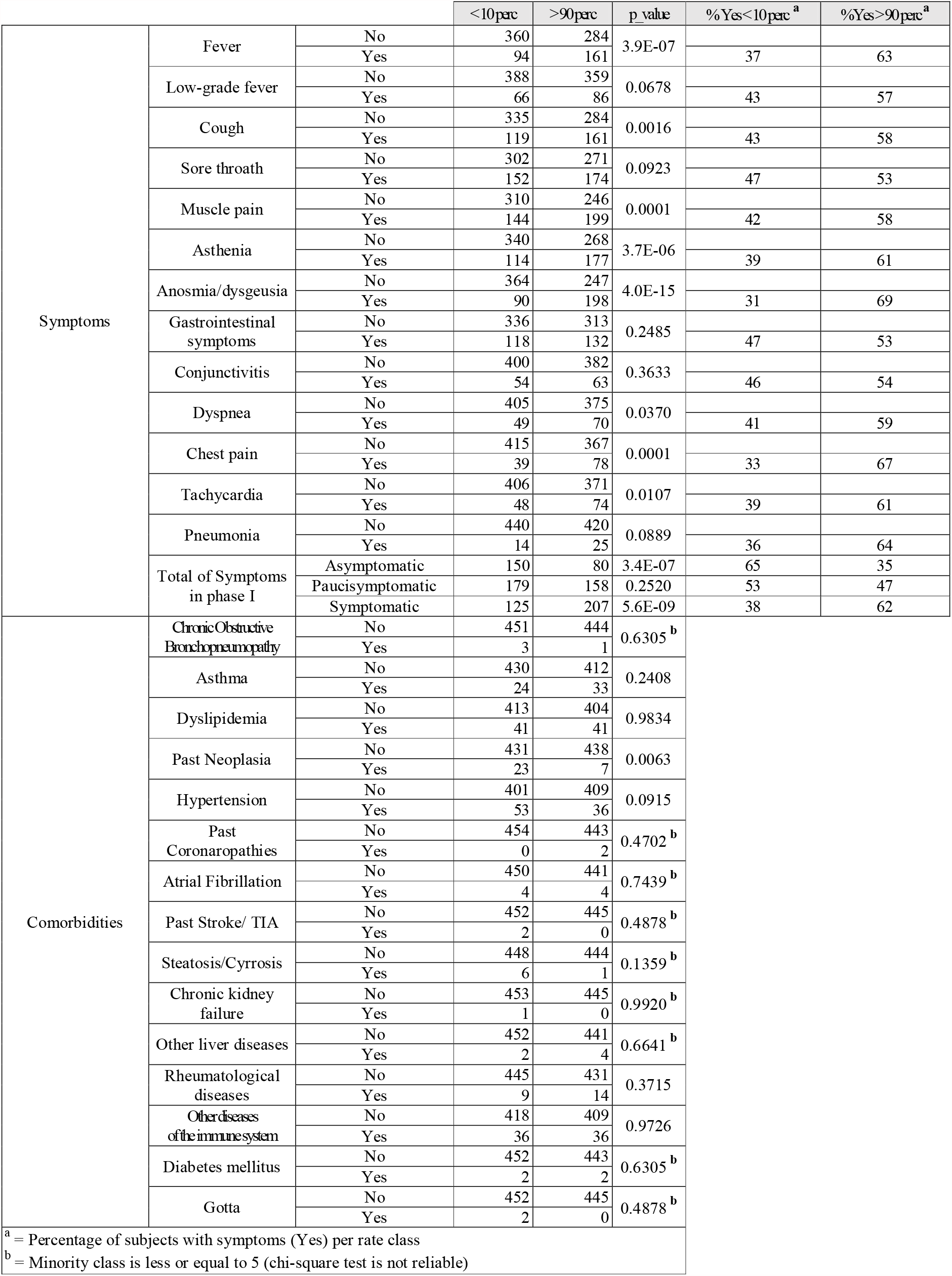
Chi-squared analysis of groups < 10^th^ percentile and > 90^th^ percentile per symptoms.

We then analyzed whether the antibody response was maintained over time in the third time point of analysis (n = 499) which was evaluated between November and December 2020 thus reaching an observation of 8-10 months. As shown in Figure 2 and Suppl. Fig. 2 we observed that both symptomatic and paucisymptomatic individuals still displayed a higher level of antibodies, however they did not increase between phase 2 and phase 3. By contrast, asymptomatic individuals did not increase their IgG levels over time.

**Figure 2:**
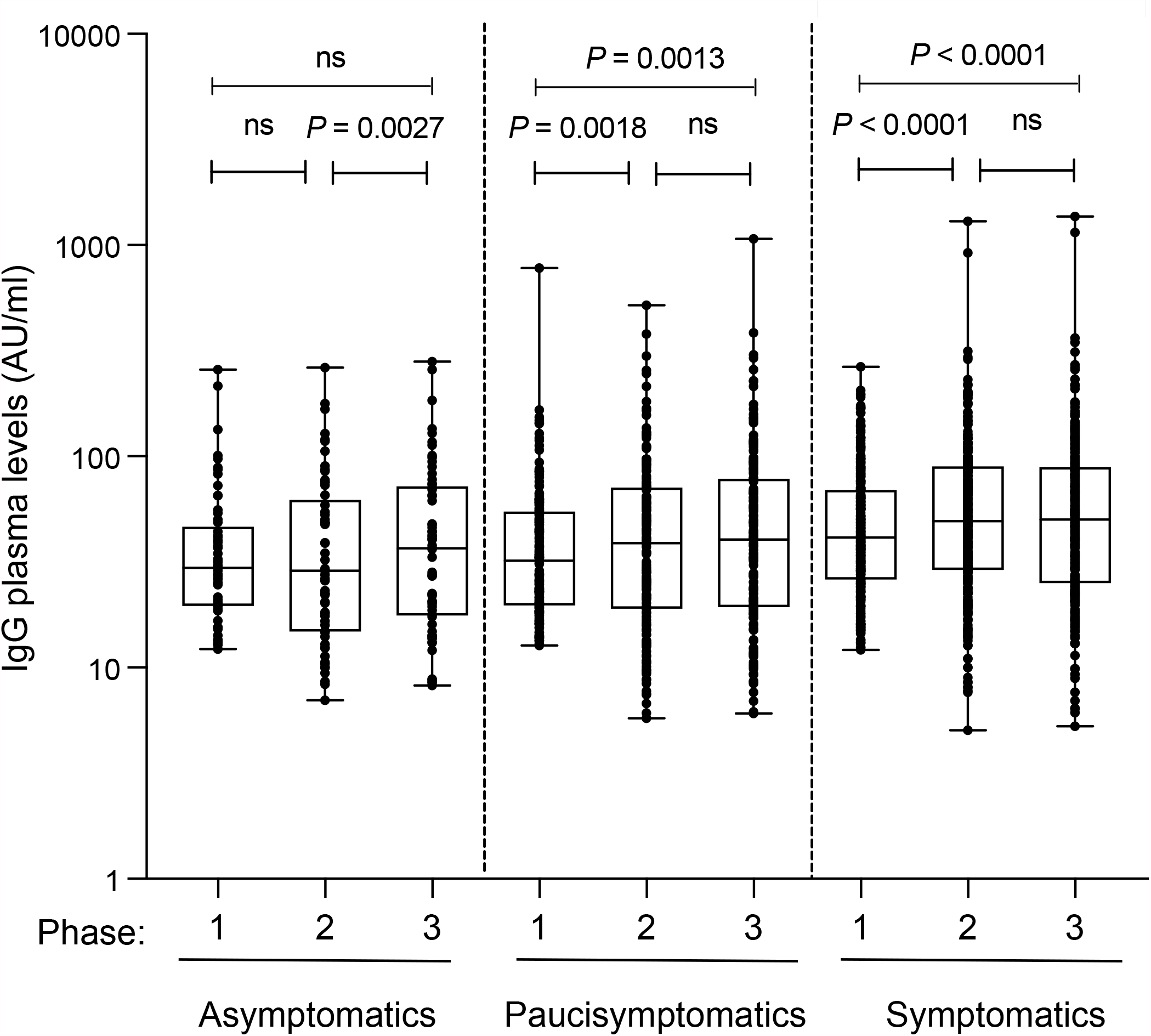
Anti-Spike S1/S2 IgG plasma levels in asymptomatics (n=61), paucisymptomatics (n=163) and symptomatics (n=275) measured at three different time points (phase 1-3). Each dot corresponds to an individual subject. Log scale on Y axis. The box plots show the interquartile range, the horizontal lines show the median values and the whiskers indicate the minimum-to-maximum range. *P* values were determined using one-tailed Wilcoxon matched-pairs signed rank test.

## Discussion

We analyzed the 8-10-month duration of an antibody response to SARS-CoV-2 in personnel from 9 healthcare facilities and an international medical school (Humanitas University) in Northern Italy in areas differently hit by the virus ^4^. We show that the antibody response is stable both in symptomatic and asymptomatic/paucisymptomatic individuals and is increased in females and in non-medical healthcare professionals. Previously, it has been shown in a study conducted in the British population that the antibody response declines of nearly 22% in symptomatic individuals and of 64% in asymptomatic individuals ^2^. However, this study was based on a prick qualitative test and thus the decline may be related to the sensitivity of the test. We also observed that the antibody response declined when we analyzed the group (3.8 < IgG < 12 AU/mL) with IgG between the limit of detection (3.8 AU/mL) and the threshold of positivity (IgG ≥ 12 AU/mL), as set by manufacturer. Whether this is linked to a difference linked to the sensitivity of the test or to a real reduction in an antibody response that may or may not be specific to SARS-CoV-2, remains to be established. In a previous analysis we excluded that this population represented individuals in the initial phases of an infection as all of them were tested for SARS-CoV-2 by a nasopharyngeal swab which however resulted negative^4^. When we analyzed the extremes, i.e. the individuals with higher rates of antibody increase or decrease (< -0.033 and > 0.005 AU/mL*day) we observed that asymptomatics had higher negative rates while symptomatics tended to continue increasing the antibody levels suggesting that extreme changes in rate separate the symptomatics from the asymptomatics. As during the observation time there was very limited viral diffusion in Northern Italy, as confirmed also by the finding that only 2 individuals became IgG positive and 2981 remained IgG negative throughout the study (all excluded from the analysis), we can conclude that the sustained or augmented antibody response may not be linked to a re-exposure to the virus. In an attempt to address what improved the antibody response, we found that several symptoms were associated to increased rates of antibodies, however, in a multivariate logistic analysis only anosmia/dysgeusia and chest pain were linked with the highest regression coefficients. Chest pain and anosmia are long-lasting symptoms in COVID-19 patients ^5^. In addition, anosmia and/or dysgeusia are very common as they are found in around 50-70% of subjects affected by COVID-19 ^6, 7^. In our cohort (Table 2), 49% of IgG positive subjects had anosmia/dysgeusia, 28% chest pain and 13.7% both anosmia/dysgeusia and chest pain, suggesting that indeed these two symptoms may, either alone or in combination, associate with IgG increase. We and others previously found that anosmia/dysgeusia together with fever were the symptoms that mostly characterized SARS-CoV-2 exposure ^4, 8^. In agreement, anosmia and dysgeusia have been proposed to be used to track SARS-CoV-2 diffusion ^9^. Interestingly, SARS-CoV-2 can infect the olfactory epithelium ^10, 11^, including olfactory sensory neurons, support cells and immune cells, that express the viral entry receptors ACE2 and TMPRSS2 ^10, 12, 13^. Here, the virus can persist long and induce local inflammation ^11^ and olfactory bulb abnormalities ^14, 15, 16^. In agreement, the loss of smell and taste can persist in individuals even with RT-PCR SARS-CoV-2 negativity in the nasopharyngeal swab for months ^11, 17^. Supporting this possibility, we did not detect any further increase of IgG levels between phase 2 and phase 3 suggesting that when individuals eliminate the virus then there is no further increase of the antibody response.

One limitation of our study is that we followed our healthcare workers for the exposure to SARS-CoV-2 via measuring the anti S1/S2 IgG response and have not evaluated any other antibody subtype nor their neutralizing activity, even though the test used has correlated the antibody levels with their neutralizing activity, as reported in the methods section.

Overall, these data suggest that increased antibody response in patients with anosmia/dysgeusia may be linked to persistence of the virus in the olfactory bulb which through local inflammation and release of antigens, maintains and boosts the antibody response. This study opens new perspectives on the immunity to SARS-CoV-2 and warrants further investigation on the role of anosmia/dysgeusia on antibody response through the design of prospective observational studies coupling the testing of SARS-CoV-2 persistence in the olfactory bulb, loss of smell or taste and antibody titers. In addition, we show that the antibody response to the natural infection is durable and persists for at least 8 months. If the antibody response elicited by the vaccines is similarly effective, we may expect it to last for at least the same amount of time. Further, this observation strongly supports our findings and those of others that convalescent symptomatic COVID-19 patients should receive only one dose of vaccine ^18, 19, 20, 21^ and suggests that this may occur even at months of distance from developing the disease as their antibody response will just need to be boosted.

## Methods

### Study population

This observational study has been approved by the international review board of Istituto Clinico Humanitas for all participating institutes (clinicaltrial.gov NCT04387929). Accrual was on a voluntary basis: it started on April 28^th^ and more than 80% of personnel participated (n = 4735). The study foresees 4 blood collections every 3/4 months. 10 different centers participate: Istituto Clinico Humanitas (ICH), Rozzano (MI); Humanitas Gavazzeni, Bergamo; Humanitas Castelli, Bergamo; Humanitas Mater Domini (HMD), Castellanza (VA); Humanitas Medical Center, HMC, Varese; Humanitas University, Pieve Emanuele (MI); Humanitas San Pio X, Milano; Humanitas Cellini, Torino; Humanitas Gradenigo, Torino; Clinica Fornaca, Torino. All participants signed an informed consent and filled a questionnaire before blood collection. We analyzed 93 features (72 categorical and 17 numerical and 4 temporal) including, age, sex, location, professional role, time between sample collections, COVID-19 symptoms (fever, sore throat, cough, muscle pain, asthenia, anosmia/dysgeusia (loss of smell and taste), gastrointestinal symptoms, conjunctivitis, dyspnea, chest pain, tachycardia, pneumonia), home exits and smart-working, comorbidities (diabetes, asthma, neoplasia, autoimmunity, cardiovascular disorders, hepatic disorders). We considered “asymptomatics” subjects without any symptoms; “paucisymptomatics” individuals that developed 1 or 2 symptoms; “symptomatics” individuals with more than 3 symptoms. None of the participants were enrolled at the time of symptoms. Thus, when the serological test was performed, they were either asymptomatics or the symptoms had disappeared. After excluding for employees that became positive for SARS-CoV-2 IgG (n = 2) during the observation period and those that dropped from phase 1 or for which we were missing at least two features, we analyzed 4534 participants (4.25% drop out). Here we show the results of the end of phase 2 and phase 3 (second and third blood sampling).

### IgG measure

For the determination of IgG anti SARS-CoV-2, the Liaison SARS-CoV-2 S1/S2 IgG assay (DiaSorin, Saluggia (VC), Italy) was used ^22^. The method is an indirect chemiluminescence immunoassay for the determination of anti-S1 and anti-S2 specific antibodies. According to kit manufacturer, the test discriminates among negative (< 15 AU/mL; with 3.8 as the limit of IgG detection) and positive (≥ 15 AU/mL) subjects. We considered positive subjects with IgG plasma levels ≥ 12 AU/mL rather than those with IgG ≥ 15 AU/mL, as suggested by the test manufacturer, based on our previous publication showing that these two groups behaved very similarly ^4^. In addition, we considered also individuals with IgG comprised between 3.8 and 12 AU/mL (which we called IgG med: 3.8 < IgG < 12 AU/mL). Consistency and reproducibility of the antibody test in samples collected in the two time points was confirmed for a limited number of individuals (n = 50) displaying different degrees of IgG positivity. The LIAISON assay’s performance in comparison to a microneutralization assay is shown in Bonelli et al. ^22^. The LIAISON serological S1/S2 assay can distinguish between neutralization positive and negative samples at cut-offs near 15 AU/mL, and additionally the data indicate that 92% of the samples with >80 AU/mL had neutralization titers ≥1:80, while 87% of samples with >80 AU/mL had neutralization titers ≥1:160.

As the samples were analyzed in separate batches, we compared the test accuracy on 21 samples from the phase 1 with the detection kits of phase 1 and phase 2 and demonstrated that the tested IgG were almost over-imposable (Suppl. Fig. 3).

### Statistical analysis and model

We first cleared the dataset by eliminating data from all of those subjects that did not develop an IgG response over time (IgG ≤ 3.8 at the beginning and at the end of the examination) (n = 2981). We then analyzed the rate of antibody response defined as:

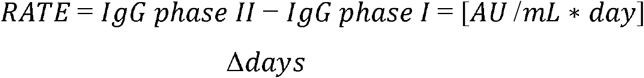

Positive rates mean increased antibody response, while negative rates indicate reduction of antibody response between the two analyzed time points.

For statistical analysis, we performed both a univariate and a multivariate analysis. We applied Wilcoxon-Mann-Whitney statistical non-parametric test to compare the antibody rate distribution between classes of subjects (Table 1 and Table 2).

We analyzed the distribution of the rate feature and found a high value of kurtosis (461) around the median value of 0.016, hence to perform a multivariate analysis we restricted the data set to subjects with IgG rates either below the 10^th^ percentile or above the 90^th^ percentile to prevent a bias-variance problem in machine learning models and subjected the data to a linear regression analysis between the training and test data sets, where the target variable (rate of antibodies) was standardized using the Yeo-Johnson method ^23^. We then applied Chi-squared statistical test to evaluate differences between classes and the rate thresholds described above (Tables 3 and 4). In order to evaluate the possible interactions between features and the rate of antibody response, we developed a multivariate approach to perform a binary classification between subjects who increased or decreased the level of antibodies. A set of 7 logistic regressions has been applied on data using a bootstrap procedure (samples are drawn with replacement) and the output of each classifier has been averaged by a Bagging classifier to obtain the final output. The selection of hyperparameters of the machine learning model and the feature selection has been performed with a Bayesian optimization approach based on cross validation (4 folds, stratified by outcome). The comparisons shown in Figure 2 and Suppl. Fig. 2 were carried out using one-tailed Wilcoxon matched-pairs signed rank test. A probability value of *P* < 0.05 was considered significant. Data analyses were carried out using GraphPad Prism version 8 and Python version 3.8 with the following libraries: Pandas (version 1.1.4, data wrangling), Scipy (version 1.3.2, statistical analysis), Scikit-Learn (version 0.24.1, LR statistical model).

## Data Availability

Humanitas metadata are deposited in Institutional Zenodo community named IRCCS Humanitas Research Hospital & Humanitas University. The dataset and the code are available at the link https://zenodo.org/record/4528974#.YCONKXnSJaQ with restricted license however available upon request.

## Acknowledgments

This work was partially supported by a philantropic donation by Dolce & Gabbana, by the Italian Ministry of Health (Ricerca corrente) and by Fondazione Humanitas per la Ricerca.

We would like to thank all the employees that volunteered to participate to this study, all the nurses and personnel that collected the samples and the laboratory technicians that run the serological and rinopharyngeal tests. We would also like to thank the Humanitas management and staff, Drs Patrizia Meroni, Michele Lagioia and Michele Tedeschi, who warmly supported this study for the safety of the employees.

## Author contribution

R.L. and L.U.: performed data analysis; C.P.: contributed to data analysis and manuscript writing; G.A. and V.S.: contributed to data analysis; M.T.S.: coordinated and supervised the laboratory analyses; E.A.: coordinated the recruitment and sampling of subjects (project administration) and participated in clinical study design; M.S.: carried out the laboratory analyses; A.M.: conceptualization and funding acquisition; M.R.: conceived the study, analyzed the data and wrote the manuscript.

