## Supplementary Figures and Tables for "The antibody response to SARS-CoV-2 infection persists over at least 8 months in symptomatic patients"

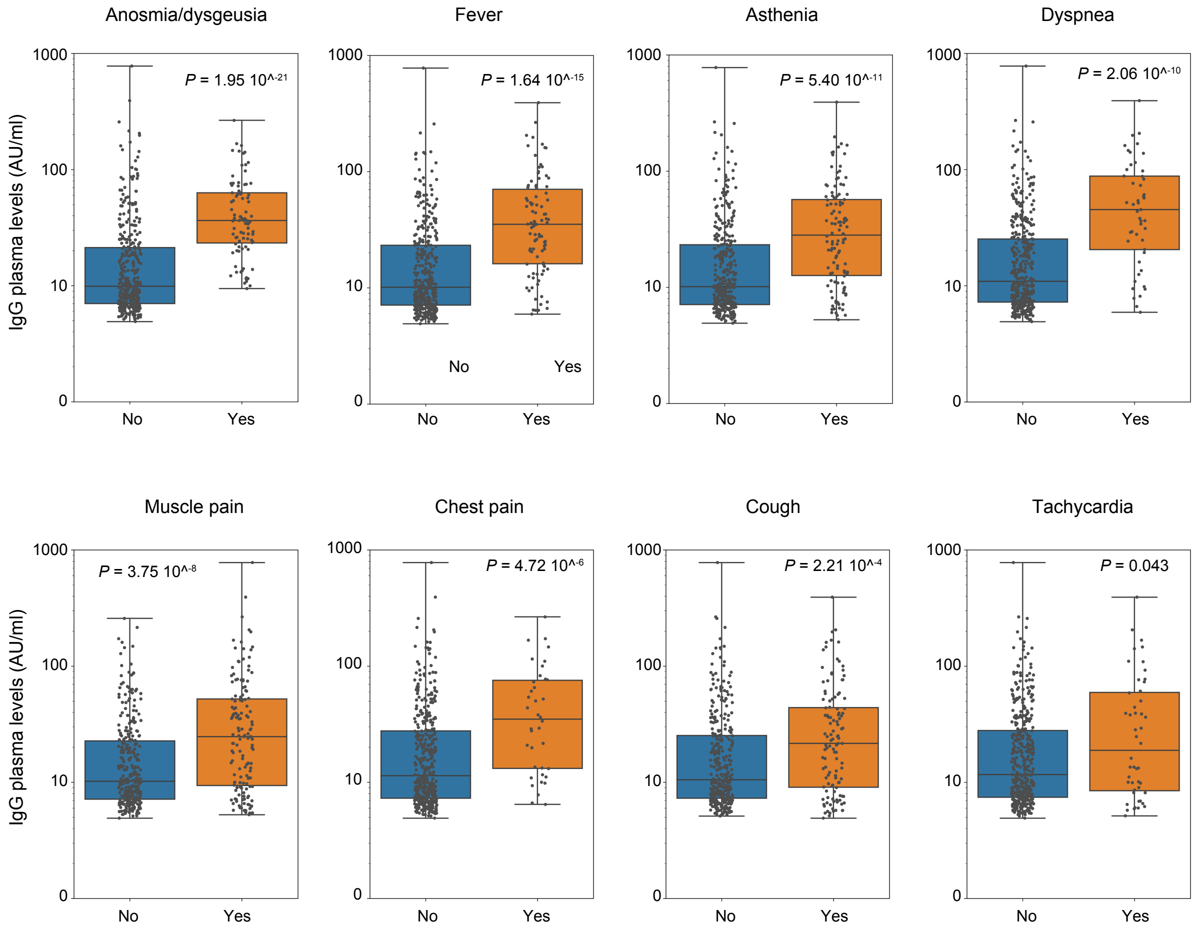


**Supplementary Figure 1.** Boxplot showing Anti-Spike S1/S2 IgG plasma levels for statistically significant symptoms in patients included in the regression model, with rates of antibodies >90$^{th}$ percentile and <10$^{th}$ percentile. *P*-values were determined using two-tailed Mann-Whitney U rank test for binary comparison.

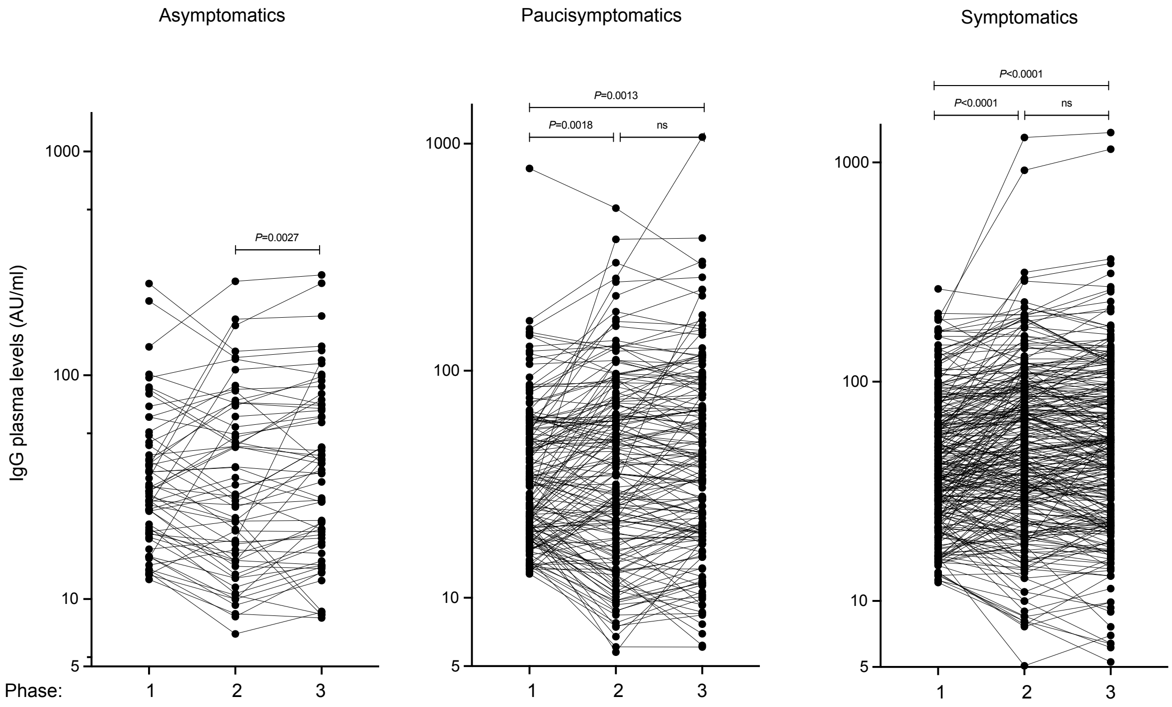


**Supplementary Figure 2.** Anti-Spike S1/S2 IgG plasma levels in asymptomatics (n=61), pauci-symptomatics (n=163) and symptomatics (n=275) measured at three different time points (phase 1-3). Connecting lines of dots correspond to the same individual in three different time points. Log scale on Y axis. *P*-values were determined using one-tailed Wilcoxon matched-pairs signed rank test.

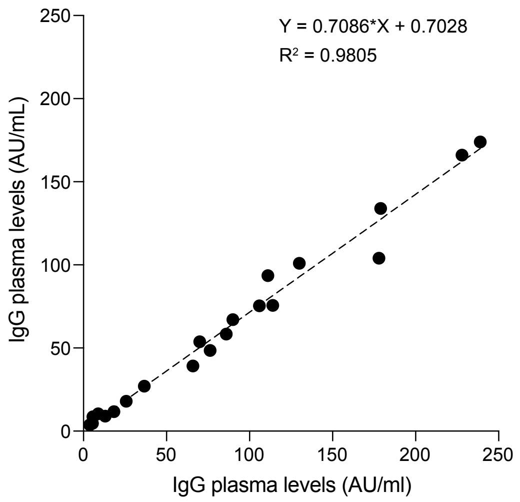


**Supplementary Figure 3.** Accuracy test on samples from the phase 1 (n=21) with the detection kits of phase 1 and phase 2.
